# Smart phone application to exclude esophageal/cardio-fundal varices in compensated cirrhosis of non-viral aetiology using liver transaminases levels and transient elastography measured liver stiffness and splenic stiffness

**DOI:** 10.1101/2022.10.17.22280110

**Authors:** AAN Nishad, MA Niriella, AP De Silva, H Jayasundara, VT Samarawickrama, H Jayasena, K Thebuwana, S Dharshika, G Hewathanthri, CK Ranawaka, M Withanage, A Pathmeswaran, HJ de Silva

## Abstract

**Introduction and objective:** We used AST to ALT ratio (AAR) and, liver stiffness measurement (LSM), splenic stiffness measurement (SSM) by transient elastography to develop a statistical model and present it as a user-friendly smartphone application to exclude the presence of oesophageal and cardio-fundal varices to avoid upper gastrointestinal endoscopy in selected patients.

**Methods:** A prospective study was carried out among patients with Child-Pugh Class A cirrhosis (non-viral and BMI<30kg/m^2^). LSM and SSM were obtained using *Fibroscan* (EchoSens) by a single operator, blinded to the presence or absence of varices. The predictors used to develop the formula were AAR, LSM and SSM. Multiple logistic regression was used to create the algorithms in 70% of the sample and validated using 30% of the sample with Bootstrapping of 1000. Best algorithms with the highest area under the curve (AUC) were selected and identified as different cut-off levels to exclude or predict the presence of varices. Those values were included in a smartphone application on android and iOS web-based platforms.

**Results:** One hundred and nine out of 211 had varices. After modelling different combinations, logistic regression formula (LRF)=5.577+(LSM*0.035)+(SSM*0.08)+(AAR*1.48) resulted AUCs 0.93. Cut-off value <-1.26 of LRF predicted the exclusion of varices with a negative predictive value of 90%. Cut-off value >0.829 of LRF predicted the presence of varices with a positive predictive value of 91%. Multiple values were used to develop a smartphone app on the Angular 2+ platform. (It can be downloaded for use @https://mediformula-65ef0.web.app/).

**Conclusion:** The new formula using AAR, LSM and SSM can be used to predict exclusion of varices with high accuracy in non-obese patients with compensated cirrhosis of non-viral aetiology based on the patient’s biochemical or fibroscan values. The smartphone application derived from this model is easy to use. It is the first mobile application to be used to exclude or predict the presence of varices utilizing SSM.

## Introduction

“Compensated advanced chronic liver disease” (cACLD) is defined as a spectrum of advanced liver fibrosis or cirrhosis in asymptomatic patients [1, 2]. Clinically significant portal hypertension (CSPH) is defined by the hepatic venous pressure gradient (HVPG) of ≥10 mmHg and is present in around 60% of patients with cACLD and in almost all patients with decompensated cirrhosis [3]. CSPH influences the development of portosystemic collaterals, leading to the development of oesophageal (EV), cardio-fundal and/or ectopic varices and is independently associated with an increased risk of decompensation and the development of hepatocellular carcinoma (HCC) [4]. Reducing the HVPG by 20% and/or to <10 mmHg significantly reduces the decompensation risk in patients with cirrhosis [5]. The gold standard for assessing portal hypertension (PH) and identifying CSPH is HVPG, which represents the gradient between the pressure in the hepatic sinusoidal capillary network and the free hepatic venous (systemic) pressure. However, owing to the limited availability of HVPG measurement, CSPH is most often diagnosed only after the detection of varices on upper-gastrointestinal endoscopy (UGIE). Despite its ease of access, endoscopy remains an invasive procedure requiring clinically trained specialists and specialized equipment. Additionally, it is generally not well perceived by patients. Furthermore, there is considerable inter-observer disagreement during the endoscopic assessment of variceal grading. [6]

Transient elastography (TE) is a non-invasive, reproducible and simple screening method which assesses tissue elasticity and, thereby, liver stiffness due to liver fibrosis [7, 11]. TE is a one-dimensional shear wave elastography (SWE) method that measures liver stiffness (LSM) or spleen stiffness (SSM) at a depth of 2.5-6.5 cm beneath the skin, with an exploration volume of 3 cm^2^ [7]. Results measured by SWE are usually presented as either m/s (tissue velocity) or kPa (estimated tissue elasticity).

In recent times, there has been extensive research investigating the potential and efficacy of LSM as a non-invasive method for diagnosing CSPH, the presence of EV and varices needing treatment (VNT). The Baveno-VII consensus statement defined non-invasive criteria based on LSM and SSM by TE and platelet count by which patients can safely avoid screening endoscopy [2]. Baveno-VII guidelines considered LSM <20kPa with platelet count more than 150,000/cm3 to “rule out” CSPH and SSM of <21kPa to “rule out” CSPH and >50 to “rule in” CSPH. [2]

In studies focusing on ruling in the presence of EV, positive-predictive values (PPV)> 90% were reported when cut-offs range from 15 kPa to 28kPa. In contrast, in studies focusing on ruling out EV, negative-predictive values (NPV)> 90% were reported when cut-offs range between 19 kPa and 48 kPa. [8, 9, 10, 11]

The use of SSM, in addition to LSM, as a non-invasive surrogate parameter for the prediction of varices, has steadily been gaining traction since it was first reported in 2013 [12]. Indeed, several studies have subsequently investigated the predictive value of SSM using TE. Notably, SSM can also capture PH secondary to both pre-sinusoidal and/or pre-hepatic causes, which may not be detected by LSM alone. Furthermore, SSM has universally been shown to be at least equal, if not superior, to LSM with regards to the detection of varices.

Aspartate aminotransferase (AST) to alanine aminotransferase (ALT) ratio (AAR) is a useful non-invasive, simple serum marker for liver cirrhosis. It is also has been successfully used to predict the presence of EV. [13, 14, 15]

Currently, there appear to be no studies evaluating the utilization of composite scores using LSM, SSM and AAR [37]. Therefore, we aimed to combine LSM, SSM and AAR to predict or exclude CSPH and the presence or absence of oesophageal and/or cardio-fundal varices. We used LSM, SSM and AAR to develop a statistical model to predict varices and present it as a user-friendly smartphone application.

## Methods

A prospective study was carried out among Child-Pugh Class A cirrhotic patients 18 to 65 years of age attending the liver clinic at the Colombo North Teaching Hospital, Ragama, Sri Lanka. The patients recruited were those with cirrhosis due to NAFLD, autoimmune or chronic unsafe alcohol use and with a BMI<30kg/m^2^. Those with cirrhosis secondary to viral hepatitis were excluded from the study. All the participants were subjected to a gastroscopy and a fibroscan.

LSM and SSM were obtained using *FibroScan® 502* Touch machine (Echosens, France) by a single operator who was blinded to the presence or absence of gastro-oesophagal or cardio-fundal varices and endoscopist was blinded for the fibroscan findings. After collecting the data 70% of the sample was randomly allocated for model building to predict the presence or absence of varices taking the gastroscopy findings as the gold standard. The predictors used for the development of the model were 1). AAR, 2). LSM and 3). SSM. Initially independent sample t-test was carried out with the predictors. Multiple logistic regression was used to develop the algorithm.

The new algorithm was internally validated using the rest (30%) of the sample with bootstrapping over 1000 [16,17,18]. The algorithm values were plotted in a ROC curve against presence or absence of varices and negative prediction value (exclusion of varices) and positive predictive values were calculated for different points at the curve. (Figure 2)

**Figure 1.**
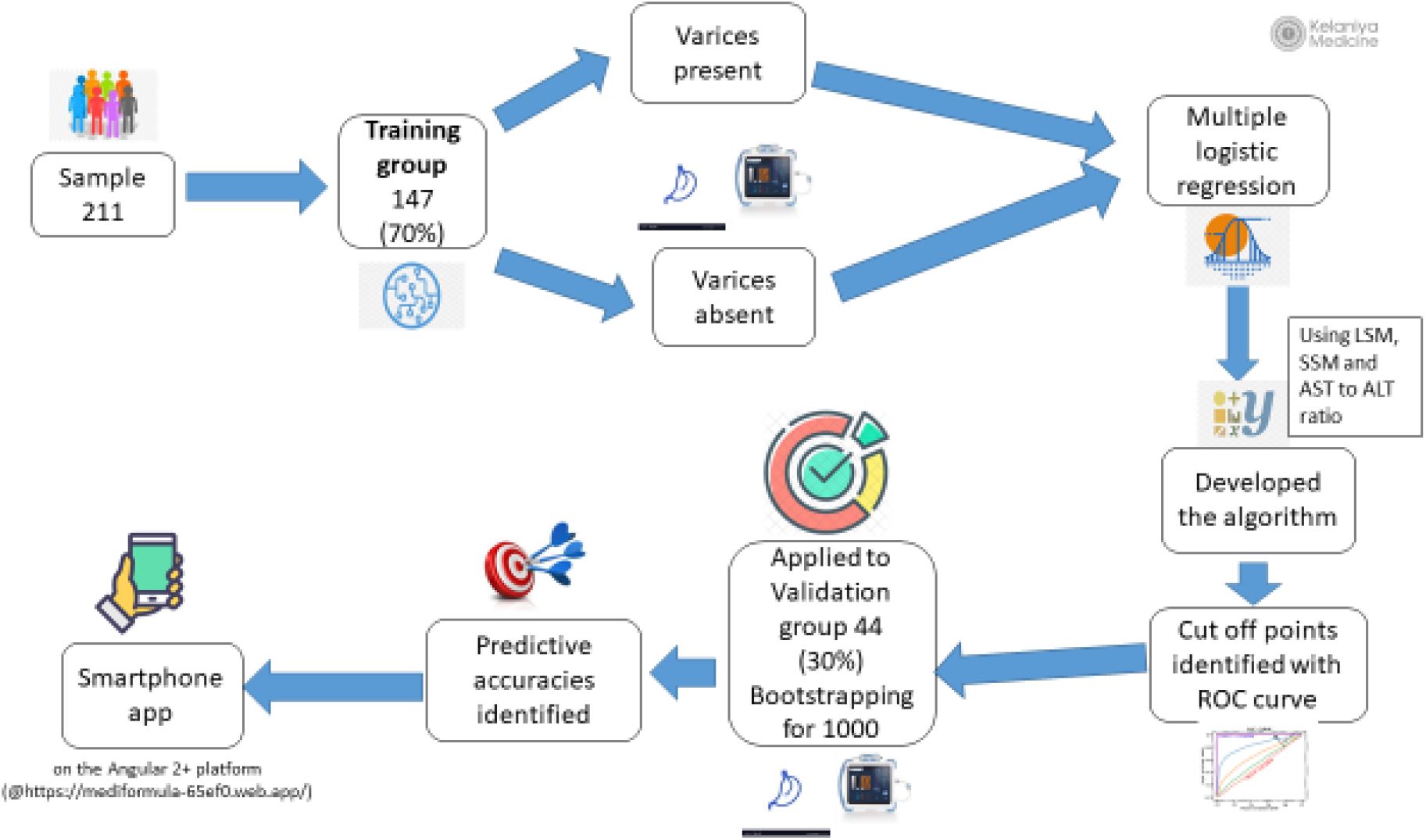
Study outline in a graphical presentation

**Figure 2.**
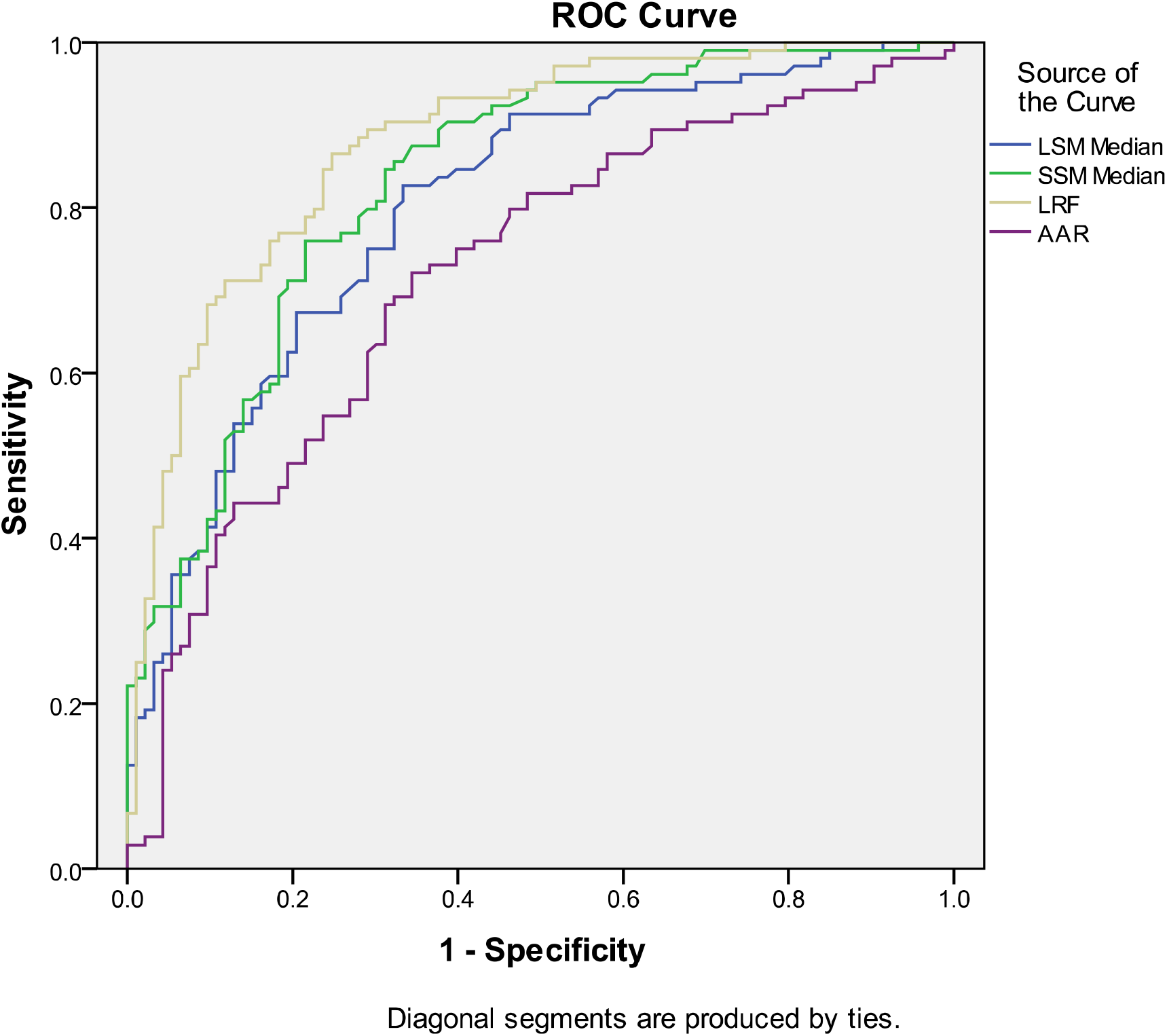
ROC curve comparing different predictors for presence of esophageal varices LRF; Logistic regression formula, LSM; Liver stiffness measurement, SSM; Spleen stiffness measurement, AAR; Aspartate aminotransferase to Alanine aminotransferase ratio

Independent sample t-tests of logistic regression formula (LRF) value were also used to cross-validate the two sub-samples for patients with varices and without varices [19, 20]. (Figure 1) Those cut of values were used to predict the varices used to build a smartphone app on the Angular 2+ platform. (It can be downloaded for use @https://mediformula-65ef0.web.app/). (Figure 3) The user needs to enter the values of LSM, SSM, AST, and ALT into the input interphase, and the app will calculate a score based on the coefficient of the logistic regression model. Then, it would identify in which range the LRF value will lie and calculate the risk of having varices or the possibility of excluding varices as its output.

**Figure 3.**
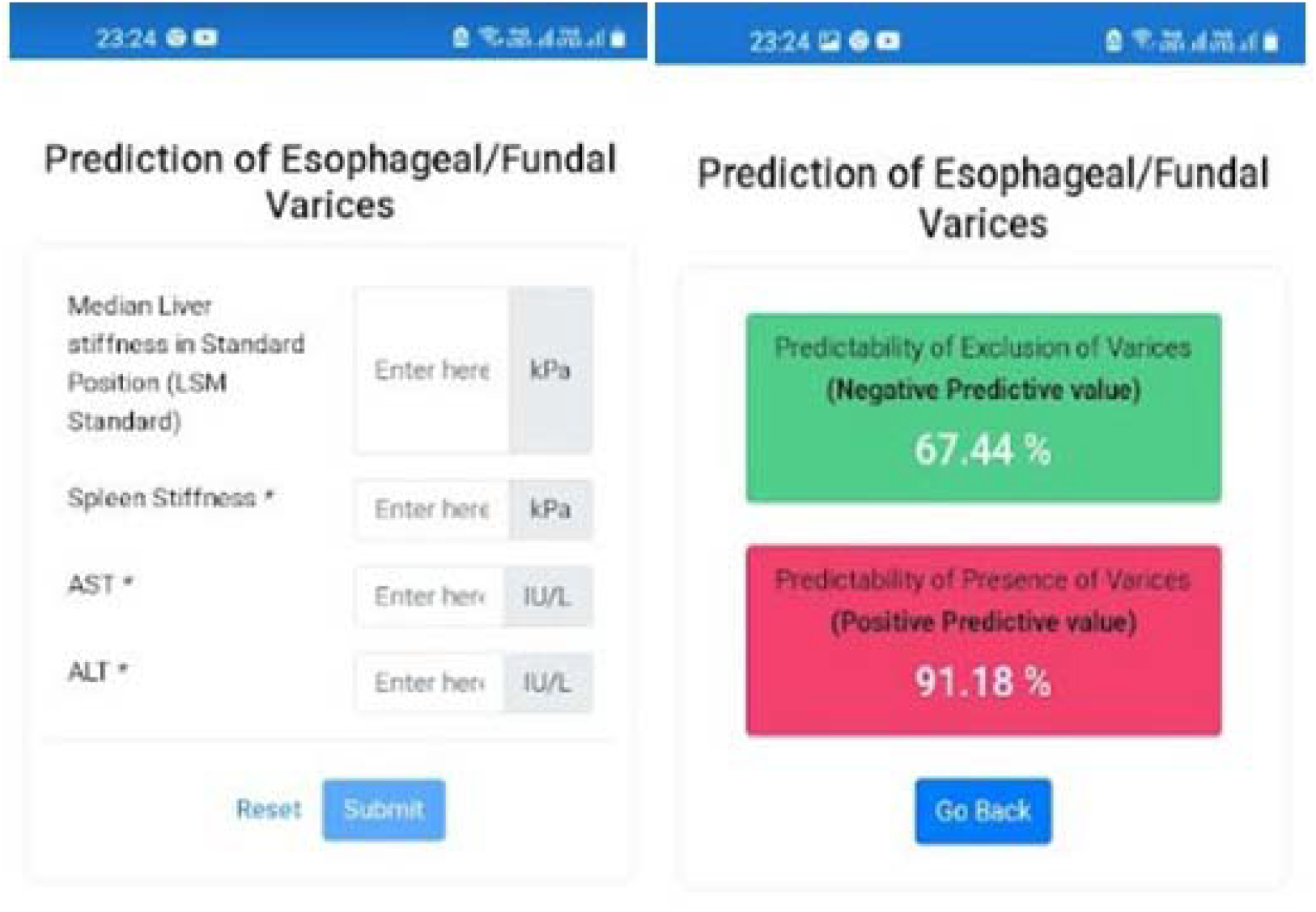
Input and output interphases of mobile app

## Results

There were 211 subjects who participated in this study. We divided the sample into two groups: training and validation. The training sample (training group) and validation sample (validation group) consisted of 154 and 67 participants respectively. The following table (Table 1) describes the socio-demographic properties of the two groups.

**Table 1.** Socio-demographic characteristics of the participants.

|  | **Training**<br>**group**<br>**(n=154)** | **Validation**<br>**group**<br>**(n=67)** | **Sig.** |
| --- | --- | --- | --- |
| BMI (Mean/s.d.)<br>kg/m<sup>2</sup> | 26.1(3.7) | 25.5(3.4) | 0.28 |
| <u>Gender</u> |  |  |  |
| Male | 58.4% | 61.2% | 0.7 |
| <u>Occupation –</u> |  |  |  |
| Currently Employed | 28.5% | 24.2% | 0.19 |
| <u>Marital status</u> |  |  |  |
| Ever Married | 92% | 97% | 0.37 |

Table 2 describes the variables used to differentiate the presence of varices vs absence of varices in the two groups the training group and the validation group.

**Table 2.** Comparison of predictive variables among training group vs validation group.

| Parameter |  | Varices absent |  |  |  | Varices present |  |  |  |
| --- | --- | --- | --- | --- | --- | --- | --- | --- | --- |
|  |  | N | Mean | 95% CI | Sig. | N | Mean | 95%CI | Sig. |
| LSM<br>Median | Training group | 67 | 20.3 | 17.5-23.1 | 0.3 | 78 | 35 | 31.2- 38.8 | 0.4 |
|  | Validation group | 32 | 17.7 | 14.1-21.3 |  | 30 | 38.3 | 30.7-45.9 |  |
| SSM<br>Median | Training group | 67 | 27.4 | 24.0-30.8 | 0.9 | 77 | 50.7 | 46.5-54.9 | 0.5 |
|  | Validation group | 33 | 27.9 | 21.7-34.1 |  | 30 | 48 | 41.6-54.4 |  |
| AAR | Training group | 67 | 1.1 | 1.0-1.2 | 0.5 | 78 | 1.4 | 1.3-1.5 | 0.7 |
|  | Validation group | 29 | 1 | 0.85-1.1 |  | 28 | 1.4 | 1.3-1.5 |  |

Those two groups did not show a statistically significant difference when comparing the presence of varices with the absence of varices, confirming that the two groups are random in allocation. When comparing the LSM, SSM and AAR in the training group all the parameters showed a statistically significant difference between the subgroup with varices and without varices as demonstrated in Table 3.

**Table 3.**
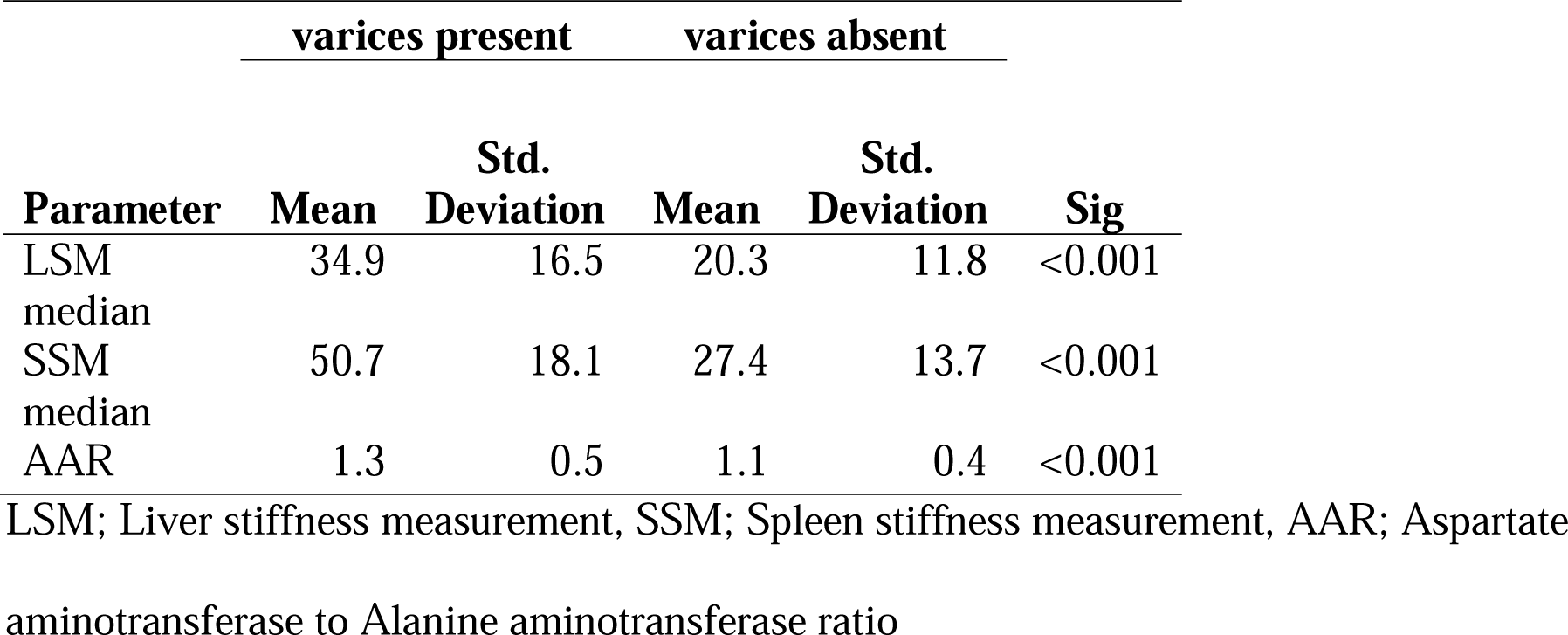
Comparison of predictive variables among the presence of varices vs absence of varices.

The mean LSM, SSM and AAR were 34.9 kPa, 50.7kPa and 1.3 respectively in those patients with varices and 20.3 kPa, 27.4 kPa and 1.1 respectively among those patients without varices. The significance values were <0.001 in all the parameters. LSM, SSM and AAR demonstrated a statistically significant difference among patients with oesophagal varices vs those without varices.

After modelling with different statistical methods, logistic regression and discriminant function analysis, the following formula value gave the highest AUC value of 0.93.

After modelling different combinations,

**Logistic regression formula (LRF) = 5.577+ (LSM*0.035)+(SSM*0.08)+(AAR*1.48)**.

(LSM; Liver stiffness measurement, SSM; Spleen stiffness measurement, AAR; Aspartate aminotransferase to Alanine aminotransferase ratio)

Area under the Curves; LRF 0.93, SSM 0.87, LSM 0.82, AAR 0.75

The following table (Table 5) shows the mean comparison of patients with varices and without varices in the modelling group vs the validation group.

**Table 4.** Validation of LRF using independent sample t-test.

|  | **LRF**<br>**value**<br>**(Mean)** | **95% CI** | **Significance** |
| --- | --- | --- | --- |
| With varices |  |  |  |
| Training group | 1.7 | 1.29-2.19 | 0.92 |
| Validation group | 1.7 | 0.85-2.45 |  |
| Without varices |  |  |  |
| Training group | -1.1 | -1.5 to -0.7 | 0.74 |
| Validation group | -1.2 | - 1.82 to -0.62 |  |

**Table 5.**
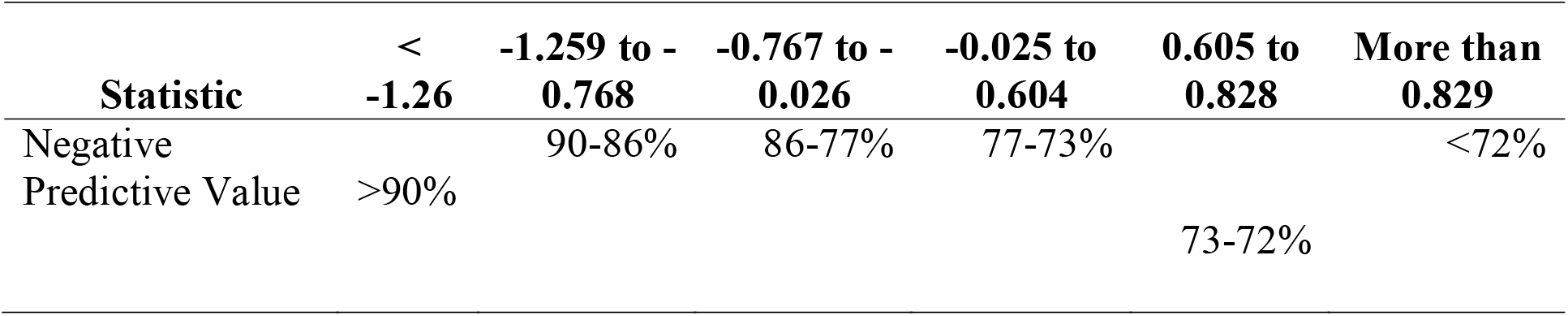
Predictive accuracies of exclusion of varices (NPV) at identified different cut of values.

The best ROC curve formula was then validated, and different cut-off values or exclusion of varices were decided and presented it as a user-friendly mobile phone application.

There is no statistically significant difference in the two groups when comparing the varices group with no varices group using independent sample t-tests.

Validation was carried out with a comparison of mean values of calculated LRF for the training group and validation group. LRF values in patients with varices in the training group and validation group were 1.7 in both groups. (p=0.92) and those values among non-varices patients were – 1.1 and -1.2 respectively. (p=0.74). (Table 4)

Cut-off value < -1.26 of LRF predicted exclusion of varices with a negative predictive value of 90%. Cut-off value > 0.829 of LRF predicted the presence of varices with a positive predictive value of 91%.

Table 5 shows that the lower the value, the higher the predictability of exclusion of varices and vice versa.

Multiple values were used to develop a smartphone app on the Angular 2+ platform.

## Discussion

We have compared few already known non-invasive markers such as LSM, SSM and AAR to predict the presence or exclusion of varices. Development of a formula and validation for exclusion of varices and presenting it as a simple user-friendly web/mobile application is the first in this subject area.

LSM, SSM and AAR were higher in patients with varices in comparison to patients without varices. (Table 3) These results are in keeping with the findings of previously reported studies [12,**Error! Bookmark not defined**.,**Error! Bookmark not defined**.,15]. LSM and SSM values are higher in patients with decompensation or in patients with varices. [21] Since all the variables are significant in mean comparison, all variables were taken into the LRF. Interestingly, the LRF with the platelet count included, gave a lower AUC. LSM, SSM and AAR gave better predictive values. Therefore, we are presenting the results without the platelet count in the prediction formula.

Even though sensitivity, specificity and accuracy are important, we have paid more attention to the ***predictive ability of the formula to exclude the presence of varices***, in order to avoid unnecessary UGIE and conversely, to determine the predictive ability to identify the presence of varices thus those patients who should be referred for a UGIE. Similar types of studies that tried to evaluate predictive accuracies have shown similar findings with different formulae and models. [22,23] Predictive accuracies for exclusion of varices (NPV) and presence of varices (PPV) of LRF values at different cut-off values were identified, so the user can decide or can inform the patient on the importance of conducting a UGIE at the given point. (Table 5, Figure 3). Coming up with a range of different cut-off values in our study made it difficult to compare with the previous studies which have given only a single cut-off or a single range and it is one of main strengths of our study. [21-38]

In the present study the cut-off value < -1.26 of LRF predicted exclusion of varices with a negative predictive value of more than 90%. In a large meta-analysis (n= 3364), demonstrated the isolated LSM cut-off of 20 kPa (alone) to predict the presence of EV with a PPV of 43% and a NPV of 86% [24]. Notably, another meta-analysis (n = 2697) reported similar results with a sensitivity of 84% and a specificity of 68% (PPV and NPV were not reported) [25].

Previous studies have demonstrated data on the combination algorithm of LSM and platelet count (commonly at the cutoff 150G/L) to rule-out VNT. But ‘Anticipate study’ results for this algorithm were rather disappointing [26]. Maurice et al evaluated these criteria in 310 patients and reported a PPV of 6% and a NPV of 98%, indicating that these criteria be highly accurate for ruling out VNT as intended [27]. Concurrently, Wong et al prospectively analyzed 274 patients and found similar results with a PPV of 9.5% and a NPV of 95.5%[28]. Moreover, a large meta-analysis by Marot including 3364 patients with mixed etiologies of liver disease reported an excellent NPV for ruling out VNT (98%) using the cut-offs as proposed by the Baveno-VI consensus [29].

The main limitation of the study is the population we used. It was limited to cirrhosis due to non-viral etiologies because we have a small number of cirrhosis due to viral diseases in Sri Lanka. Therefore, the generalizability of the findings of this study to all cirrhotics may be limited. But interested researchers can use our methodologies to explore the possibility of applying the same principles to those patients as well. We also have studied the platelet count in addition to LSM and SSM as suggested by the Baveno 7 criteria. It has not been shown in the results because of its low sensitivity, specificity, NPV and PPV in comparison to our new formula.

## Conclusion

The new formula using AAR, LSM and SSM can predict varices with high accuracy in non-obese patients with compensated cirrhosis of non-viral aetiology. The smartphone application derived from this model is user-friendly, and it is the first mobile application of its kind again proving the comment by the review in 2015. [**Error! Bookmark not defined**.]

### Images

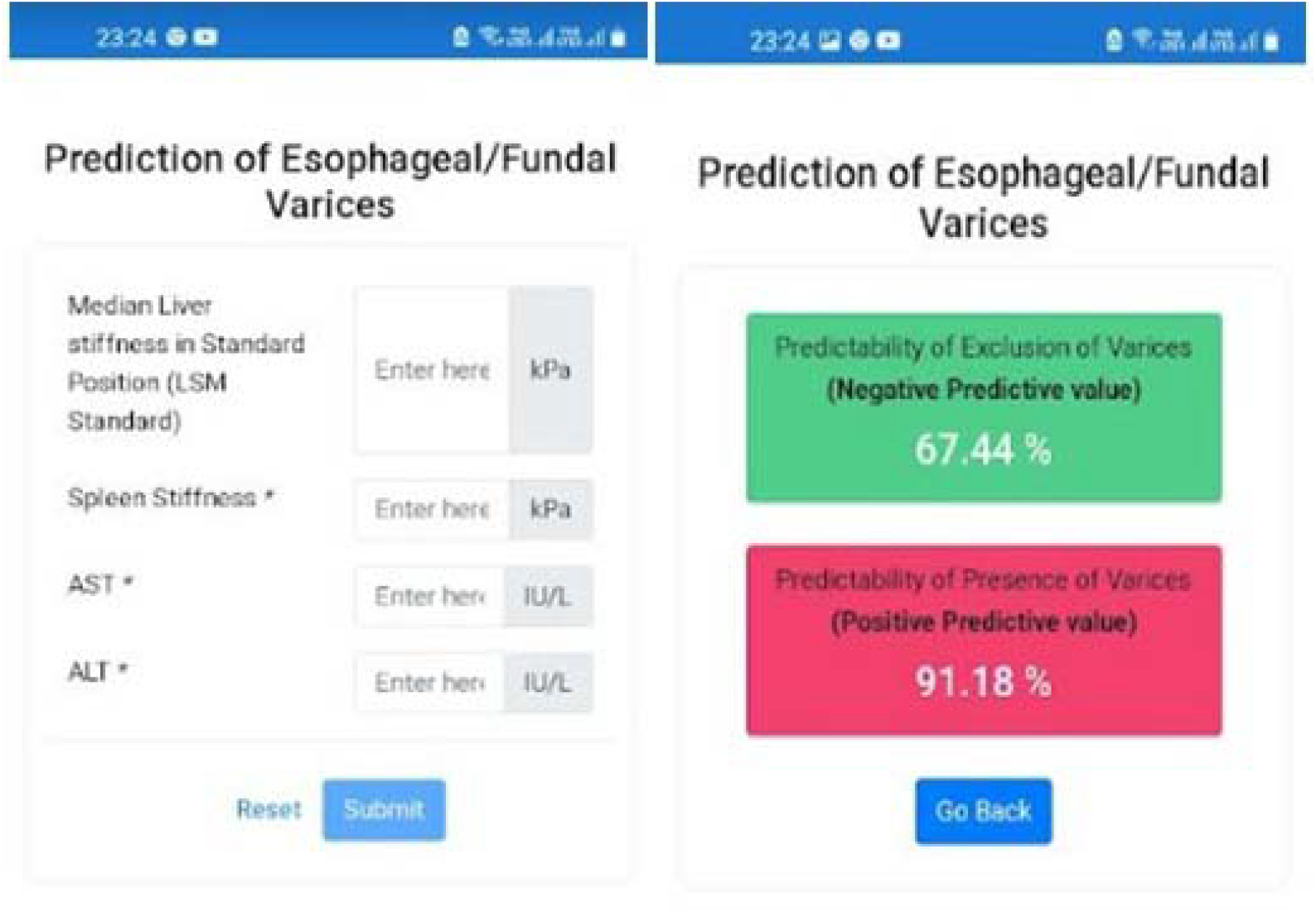

## Data Availability

All data produced in the present work are contained in the manuscript

